# The STRAT Clinical Risk Score to Predict Early Ischemic Stroke Post-TAVI: findings from the FRANCE-TAVI registry

**DOI:** 10.1101/2025.08.01.25332651

**Authors:** Matthieu Besutti, Romain Chopard, Fiona Ecarnot, Manon Leclère, Hélène Eltchaninoff, Bernard Iung, Martine Gilard, Philippe Commeau, Mariama Akodad, Hakim Benamer, Sylvain Beurtheret, Thomas Cuisset, Hervé Le Breton, Eric Van Belle, Jean-Philippe Verhoye, Guillaume Cayla, François Schiele, Maxime Desmarets, Nicolas Meneveau, the RHU STOP-AS and France-TAVI Investigators

**Affiliations:** Department of Cardiology, University Hospital Besançon, Besançon, France; SINERGIES, Université Marie & Louis Pasteur, 25000 Besançon, France; F-CRIN, INNOVTE network, France; Clinityx, 92100 Boulogne-Billancourt, France; Cardiology Service, Rouen-Charles-Nicolle University Hospital Center, National Institute of Health and Medical Research U644, Rouen, France; RHU STOP-AS, 76000 Rouen, France; Department of Cardiology, Bichat Hospital, AP-HP, Paris, France; Department of Cardiology, Brest University Hospital Center, Brest, France; Service de Cardiologie Interventionnelle, Polyclinique Les Fleurs, Groupe ELSAN, 83190 Ollioules, France; Institut Cardiovasculaire Paris-Sud, Hôpital Jacques Cartier, Ramsay-Santé, 91300 Massy, France; Institut Cardiovasculaire Paris Sud, Hôpital Jacques-Cartier, Ramsay Santé, 91300 Massy, France; Department of Cardiac Surgery, Saint Joseph Hospital, Marseille, France; Department of Cardiology, La Timone University Hospital Center, Public Assistance Hospitals of Marseille, National Institute of Health and Medical Research UMR1062, French National Institute for Agricultural Research UMR 1260, University of Aix-Marseille, Marseille, France; Cardiology and Vascular Diseases Service, Pontchaillou University Hospital Center, Center for Clinical Investigation 804, University of Rennes 1, Signal and Image Treatment Laboratory (LTSI), National Institute of Health and Medical Research U1099, Rennes, France; Department of Cardiology, University of Lille 2, Regional University Hospital Center of Lille, National Institute of Health and Medical Research U1011, University Hospital Federation Integra, Lille, France; Thoracic and Cardiovascular Surgery Service, Pontchaillou University Hospital Center, University of Rennes 1, Signal and Image Treatment Laboratory (LTSI), National Institute of Health and Medical Research U1099, Rennes, France; Department of Cardiology, CHU Nîmes, 30029 Nîmes, France; CHU Besançon, Centre d’investigation Clinique 1431, F-25000 Besançon, France; Université de Franche-Comté, EFS, INSERM, RIGHT (UMR 1098), F-25000 Besançon, France

**Keywords:** Ischemic Stroke, Transcatheter Aortic Valve Implantation, risk prediction score.

## Abstract

**Background:** Practitioners recommending transcatheter aortic valve implantation (TAVI) currently lack reliable tools to predict periprocedural ischemic stroke risk. We aimed to develop and internally validate a clinical risk score to accurately stratify this risk.

**Methods:** Using data from the nationwide, multicenter FRANCE-TAVI registry, we developed a clinical predictive risk score for 30-day ischemic stroke post-TAVI using multivariable logistic regression analysis. The model was internally validated through cross-validation techniques.

**Results:** Among 62,747 patients, 1,712 (2.7%), experienced ischemic stroke within 30 days. Nine clinical predictors were identified, namely female sex, age >85 years, weight <60 kg, symptomatic status, history of stroke or transient ischemic attack, multiple (i.e., >1) episodes of acute heart failure, severe mobility reduction, diabetes, and creatinine clearance <60 mL/min. The resulting scoring model demonstrated good accuracy (Brier score, 0.17), moderate discrimination (C-index, 0.63), and excellent calibration as assessed by calibration plots, calibration-in-the-large, and calibration slope. The score categorized patients into low (90.2% of the population), intermediate (8.0%) and high-risk (1.8%) groups. Observed stroke rates increased progressively across these groups, from 2.25% in the low-risk group to 6.51% in the intermediate-risk group and 10.10% in the high-risk group.

**Conclusions:** This newly developed STRAT score is a clinical, practical, and effective tool for predicting early ischemic stroke in patients undergoing TAVI. It may help tailor preventive strategies. Further studies are necessary to externally validate this score and evaluate its impact on clinical decision-making.

## INTRODUCTION

Transcatheter aortic valve Implantation (TAVI) has become the mainstream therapy for patients with severe, symptomatic aortic stenosis (AS) who are at intermediate, high, or prohibitive risk for surgical aortic valve replacement (SAVR) [1–4]. Technological and procedural improvements, together with increased operator experience, have led to progressive widening of the indications for TAVI to include younger patients, starting from 75 [5] and even 65 years old [6], who are at low surgical risk. Nevertheless, despite these improvements, ischemic stroke still represents the most unpredictable and feared complication following TAVI, with an unchanged rate of occurrence over the years [7]. Data from prospective trials report stroke incidence ranging from 0.6% to 6.7%, while contemporary real-life registries report a consistently low but seemingly irreducible average incidence of periprocedural TAVI-related stroke at approximately 2.3% [8]. Disabling stroke related to TAVI carries a three- to nine-fold increased mortality risk; 40% of survivors have moderate to severe permanent disability leading to dependence, and 80% face social isolation and significant financial strain [9–12]. Given the extensive indications for TAVI, it is of paramount importance to avail of reliable tools for stroke prediction to tailor peri- and post-procedural management in patients undergoing TAVI. Numerous potential patient- and procedure-related predictors of neurological events have been identified [8], but they have not been aggregated into a reliable scoring system. Therefore, using data from the nationwide FRANCE-TAVI registry, we derived and internally validated a clinical prediction score for ischemic stroke at 30 days in patients undergoing TAVI, identifying patients with either high or low probability of experiencing early ischemic stroke.

## METHODS

### FRANCE-TAVI registry

The present study used data from the FRANCE-TAVI registry, which has previously been described elsewhere [13]. Briefly, designed as an all-comer registry, the FRANCE-TAVI registry prospectively included all patients undergoing TAVI in 48 of the 50 active French centers from January 2013 onwards. The registry was designed to obtain baseline characteristics of the patients, as well as TAVI procedural features and outcomes. As previously described, the decision to perform TAVI, the choice of access, and the type of device were made based on assessment by a multidisciplinary heart team [13]. Procedures and postprocedural management were performed in accordance with each site’s routine protocol. The dataset was collected using a dedicated internet-based interface managed by the French Society of Cardiology, which implements regular data quality checks, including range checks and assessments of internal consistency. All patients provided written informed consent for anonymous processing of their data, and the Institutional Review Board of the French Ministry of Health approved the registry. All data are the property of the French Society of Cardiology and were collected with the participation of the French Society of Thoracic and Cardiovascular Surgery. As previously described [14], since 2019, it has been linked with the single-payer National Health Data System (SNDS) with specific authorization from the French national data privacy commission (CNIL; National Data Protection Commission), allowing comprehensive and reliable clinical follow-up over time. Further information on the linkage is available in the **supplementary material**.

### Inclusion / Exclusion criteria

All patients included in the FRANCE-TAVI registry from its inception in 2013 until the end of 2021 were considered for the analysis. We excluded from the present analysis any SNDS entries with failure of probabilistic linkage, incorrect patient data (such as wrong date of birth, incorrect admission date, multiple dates of death, or duplicate data), and patients who did not ultimately have a valve implanted. Patients with non-femoral access were also excluded, to ensure a homogeneous population.

### Outcome and definitions

The primary outcome was ischemic stroke at 30 days. In the FRANCE-TAVI registry, an ischemic stroke was defined as rapid onset of a neurological deficit related to cerebral infarction. In the SNDS, stroke is recorded in hospital discharge reports using the 10th revision of the International Classification of Diseases version 10 (ICD-10). The following codes were used for analysis: I63 (cerebral infarction); I64 (stroke, not specified as haemorrhage or infarction: this latter category is used only in the absence of diagnostic imaging, for example if the patient dies before any examination can be performed).

Symptomatic status of aortic stenosis was defined by the presence of cardiovascular functional symptoms attributed to aortic stenosis. Severe reduction in mobility could be secondary to musculoskeletal or neurological dysfunction. High-volume centers were defined as those performing at least 150 TAVI procedures per year.

### Statistical methods

#### Model construction

The entire study population was used for the development and internal validation of the predictive score, since current recommendations advise against data splitting [15]. Continuous variables are expressed as mean (standard deviation) and categorical variables as number (percentage). Unadjusted differences between patients who had an ischemic stroke and those who did not were compared using the Chi-square test or Student’s t-test, as appropriate. The full list of candidate clinical covariates is given in **Supplemental Table 1**. Missing data were handled with multiple imputations using logistic regression for binary variables and predictive mean matching for continuous variables [16]. Continuous variables that were statistically significant were categorized, choosing the most discriminative cut-off points, based on the best-subset selection. Potential covariates were selected for the multivariable logistic regression models if they were either known to be associated with ischemic stroke (e.g. diabetes [17]) or had a p-value <0.05, excluding variables with more than 20% missing data [18]. The potential for covariate multiple collinearity was tested using the variance inflation factor (VIF) and condition number (CN), with VIF <10 and CN <30 as reference values [19].

#### Risk score construction

A risk score for predicting the likelihood of ischemic stroke at 30 days was developed from the logistic regression model. The contribution of each variable to the risk score was calculated using the natural logarithm of the exp (β coefficients) obtained from the multivariable analysis. The risk score formula was constructed as follows:

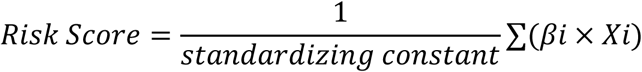

where *βi* represents the β coefficient for each predictor variable, and *Xi* is the presence (1) or absence (0) of the corresponding variable. To standardize the score, the sum of the weighted predictors was divided by a standardizing constant that corresponded to the maximum possible score. This approach ensured that the final risk score was easier to interpret (ranging between 0 and 10) [20].

#### Performance capacities

The Brier score was calculated to measure prediction accuracy, Discrimination of the model was assessed using the C-statistic, while calibration was evaluated by visually plotting the mean predicted ischemic stroke at 30 days against observed stroke in each decile. Calibration-in-the-large and Calibration Slope were used to assess overall agreement and the spread of estimated risks relative to observed outcomes, respectively [15]. The risk score was then collapsed into “low”, “intermediate” and “high” risk groups based on the observed stroke rate. We assessed the ability of the score to predict the risk of 30-day ischemic stroke by running univariable logistic regression (odds ratios [ORs] and 95% confidence intervals [CI]), with the low-risk category as the reference group vs intermediate and high-risk categories.

#### Internal validity

We validated the model internally using a K-fold cross validation. Model performance was evaluated by splitting the data into k=5 groups. In each iteration, k-1 groups were used to develop a (temporary) model, repeating the model building steps on all the data, while the remaining group was used to evaluate the performance of the temporary model. This process was repeated k times, each time leaving out a different group, producing k values for each performance measure. The performances of the developed model were then taken as the average of these k performance measures [15].

#### Sensitivity analyses

Sensitivity analyses were performed to assess the robustness of the findings. These included analyses restricted to complete cases, ICD-10 code I63, and periprocedural stroke (0-3 days), as well as stratification by treatment period and center procedure volume. An internal-external validation was also performed by dividing France into five major geographic regions [15].

#### Competing risk

To investigate whether the risk of death varied according to score levels, we analyzed death rates according to Ischemic Stroke score levels [21].

The study was performed in accordance with the recommendations of the TRIPOD and STROBE statements [22, 23]. A p-value < 0.05 was considered significant. Analyses were performed using Python version 3.8 (Python Software Foundation, Wilmington, DE).

## RESULTS

### Study population

A total of 80,620 patients were included in the FRANCE-TAVI registry from 2013 to the end of 2021. Among these, 7,551 had failure of probabilistic linkage to the SNDS, 362 had no valve implantation, 9,956 had non-femoral access. The remaining 62,747 patients composed the study population **(Figure 1).** Mean age was 82.7 ± 6.8 years; 31,310 (49.9%) were men. Mean EuroSCORE II was 5.3 ± 5.4. Symptoms were present in 59,079 (96.9%) patients and 4,721 (8.2%) experienced acute heart failure. Comorbidities included diabetes mellitus in 16,394 (26.5%), previous stroke in 6,430 (10.4%), chronic renal failure in 24,706 (41%), and atrial fibrillation in 22,421 (35.7%) patients. Severe reduction in mobility was observed in 3,966 (6.5%) patients. Regarding the type of valve, 22,520 (37.9%) patients received a self-expandable valve and 36,848 (62.1%) received a balloon-expandable valve. Of these, 86.7% were third-generation valves (**Table 1)**.

**Figure 1.**
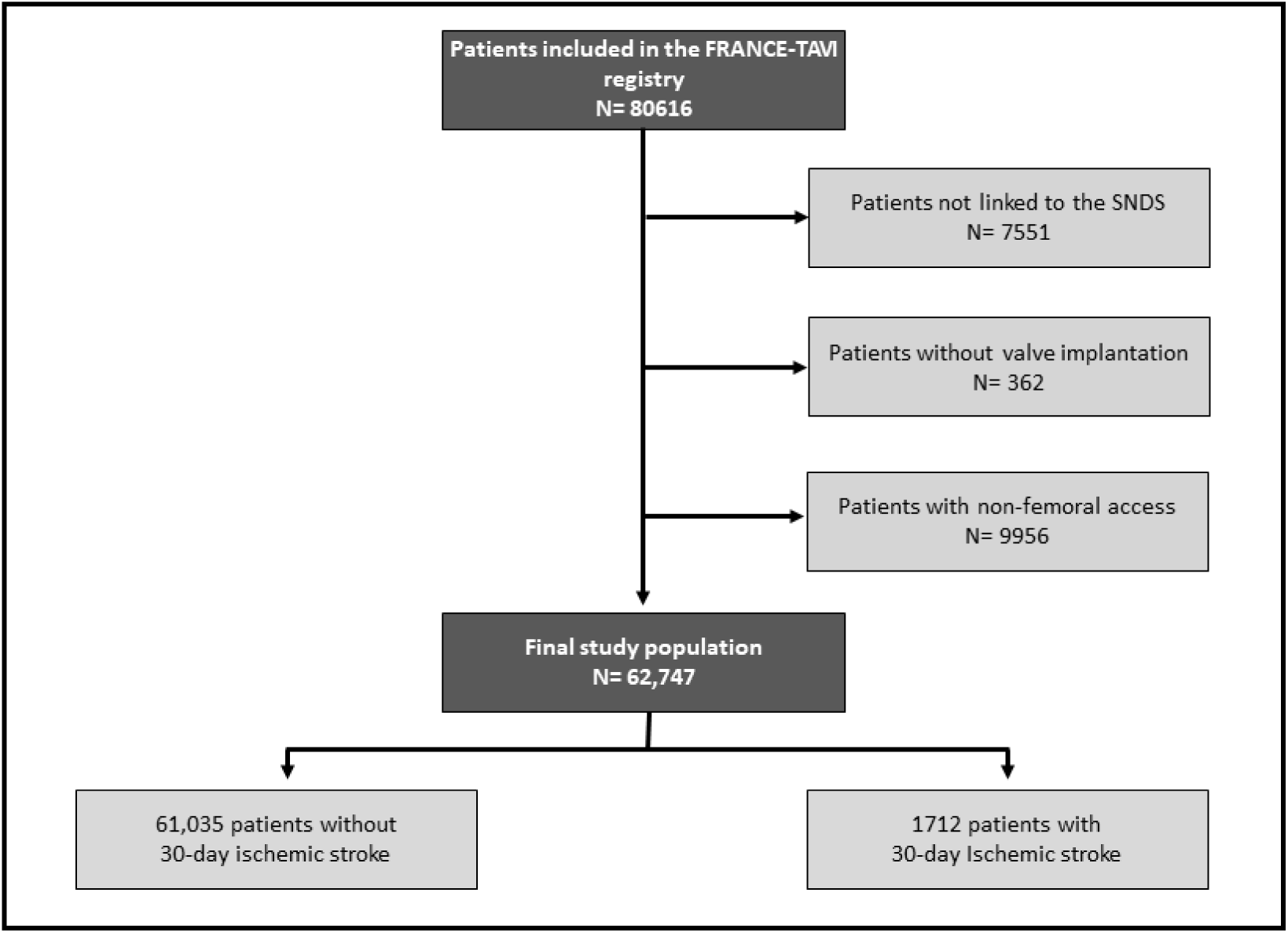
Study Flowchart. SNDS : Système National des Données de Santé (National health data system)

**Table 1.** Baseline characteristics and clinical data of 62747 study patients according to the occurrence of Ischemic Stroke (IS). **Abbreviations:** CABG: Coronary Artery Bypass Graft Surgery; COPD: Chronic Obstructive Pulmonary Disease; NYHA: New York Heart Association functional classification; PCI: Percutaneous Coronary Intervention; PAD: Peripheral artery disease; SAVR: Surgical Aortic Valve Replacement; Transient Ischemic Attack; § More than one episode of acute heart failure in the past 12 months; *SNDS data. ** Cockcroft Clearance <60 ml/min.

| Variable | Missing, n (%) | All<br>(n=62747) | No stroke at<br>30 days (n=61035) | Stroke at 30<br>days (n=1712) | p value |
| --- | --- | --- | --- | --- | --- |
| **General Information** |  |  |  |  |  |
| Age, years (sd) | 0 (0) | 82.7 ± 6.8 | 82.7 ± 6.8 | 83.7 ± 6.5 | <0.001 |
| Male sex, n (%) | 0 (0) | 31310 (49.9) | 30561 (50.1) | 749 (43.8) | <0.001 |
| BMI, kg/m2 (sd) | 0 (0) | 26.9 ± 5.2 | 26.9 ± 5.2 | 26.3 ± 5.1 | <0.001 |
| **Surgical Risk Scores** |  |  |  |  |  |
| Logistic EuroSCORE, n (%) | 5921 (9.4) | 15.6 ± 11.8 | 15.6 ± 11.8 | 17.9 ± 13.3 | <0.001 |
| EuroSCORE II, n (%) | 23616 (37.6) | 5.3 ± 5.4 | 5.3 ± 5.4 | 6.0 ± 6.2 | <0.001 |
| **Symptoms** |  |  |  |  |  |
| Symptomatic status, n (%) | 1783 (2.8) | 59079 (96.9) | 57441 (96.9) | 1638 (98.4) | 0.006 |
| NYHA III/IV, n (%) | 7818 (12.5) | 33545 (61.1) | 32613 (61.0) | 932 (62.4) | 0.28 |
| Acute Heart Failure§, n (%) | 5440 (8.6) | 4721 (8.2) | 4556 (8.2) | 165 (10.7) | <0.001 |
| **CV Risk Factors** |  |  |  |  |  |
| Diabetes mellitus, n (%) | 881 (1.4) | 16394 (26.5) | 15921 (26.5) | 473 (28.2) | 0.125 |
| Arterial Hypertension*, n (%) | 0 (0) | 42964 (68.5) | 41750 (68.4) | 1214 (70.9) | 0.03 |
| Dyslipidemia*, n (%) | 0 (0) | 16222 (25.9) | 15741 (25.8) | 481 (28.1) | 0.03 |
| **Comorbidities** |  |  |  |  |  |
| Previous SAVR, n (%) | 1209 (1.9) | 275 (< 1%) | 268 (< 1%) | <11 (< 1%) | 1.00 |
| Previous CABG, n (%) | 60 (<1) | 575 (< 1%) | 561 (< 1%) | 14 (< 1%) | 0.75 |
| Previous PCI, n (%) | 841 (1.3) | 17782 (28.7) | 17309 (28.7) | 473 (28.2) | 0.66 |
| Previous stroke/TIA, n (%) | 861 (1.4) | 6430 (10.4) | 5991 (10.0) | 439 (26.1) | <0.001 |
| PAD, n (%) | 321 (19.1) | 10267 (16.6) | 9946 (16.5) | 148 (8.9) | 0.006 |
| Chronic renal failure**, n (%) | 2410 (3.8) | 24706 (41) | 23950 (40.8) | 576 (46.4) | <0.001 |
| Cockcroft Clearance | 4595 (7.3) | 52.4 ± 22.5 | 52.5 ± 22.5 | 48.2 ± 21.0 | <0.001 |
| COPD*, n (%) | 0 (0) | 7766 (12.4) | 7553 (12.4) | 213 (12.4) | 0.96 |
| Atrial fibrillation*, n (%) | 0 (0) | 22421 (35.7) | 21757 (35.6) | 664 (38.8) | 0.008 |
| Permanent Pacemaker, n (%) | 851 (1.4) | 7119 (11.5) | 6942 (11.5) | 177 (10.6) | 0.23 |
| Severe reduction in mobility, n (%) | 1393 (2.2) | 3966 (6.5) | 3815 (6.4) | 151 (9.1) | <0.001 |
| **Echocardiography at Baseline** |  |  |  |  |  |
| Ejection fraction, n (%) | 3300 (5.3) | 55.5 ± 14.4 | 55.5 ± 14.4 | 55.5 ± 14.4 | 0.94 |
| Aortic valve area, cm2 (sd) | 12923 (20.6) | 0.73 ± 0.24 | 0.7 ± 0.3 | 0.7 ± 0.2 | 0.001 |
| Mean gradient, mmHg (sd) | 6533 (10.4) | 47.9 ± 15.0 | 47.8 ± 15.0 | 48.2 ± 15.4 | 0.76 |
| Aortic regurgitation ≥ grade 2, n (%) | 16050 (25.6) | 9527 (20.4) | 9282 (20.4) | 245 (19.8) | 0.59 |
| Mitral regurgitation ≥ grade 2, n (%) | 17179 (27.4) | 9707 (21.3) | 9446 (21.3) | 261 (21.3) | 1.00 |
| sPAP > 60 mmHg, n (%) | 17104 (27.3) | 4437 (9.7) | 4306 (9.7) | 131 (10.6) | 0.33 |
| **Procedure** |  |  |  |  |  |
| Type of valve, n (%) | 3379 (5.4) |  |  |  | <0.001 |
| - Self-expandable |  | 22520 (37.9) | 21714 (96.4) | 806 (3.6) |  |
| - Balloon-expandable |  | 36848 (62.1) | 36043 (97.8) | 805 (2.2) |  |
| Need for a second valve, n (%) |  | 849 (1.4) | 781 (1.3) | 68 (4.0) | <0.001 |
| **Echocardiography at Discharge** |  |  |  |  |  |
| Ejection fraction, n (%) | 11289 (18.0) | 51.3 ± 21.3 | 51.3 ± 21.3 | 51.1 ± 21.1 | 0.29 |
| Aortic valve area, cm2 (sd) | 43222 (68.9) | 1.9 ± 0.6 | 1.8 ± 0.6 | 1.8 ± 0.6 | 0.06 |
| Mean gradient, mmHg (sd) | 14149 (22.5) | 10.5 ± 5.8 | 10.5 ± 5.8 | 9.7 ± 5.8 | <0.001 |
| Aortic regurgitation ≥ grade 2, n (%) | 14517 (23.1) | 19778 (41.0) | 19221 (40.9) | 557 (44.4) | <0.001 |

### Outcomes

Data for the primary outcome of ischemic stroke at 30 days was complete, with no missing values. Overall, 1,712 (2.7%; 95% CI: 2.57%, 2.82%) experienced an ischemic stroke within 30 days post-procedure (of which 10.9% ICD-10-defined I64 stroke related death) with a median time to event of 0 days (Q1-Q3, 0-7 days, ranging from 0 to 30 days). The remaining 61,035 (97.3%; 95% CI: 97.18%, 97.43%) patients did not experience stroke. Most strokes occurred in the peri-procedural period: 73.4% (95% CI: 71.3%, 75.5%) within the first 3 days post-TAVI, with the vast majority occurring during the procedure, while 26.5% (95% CI: 24.5%, 28.5%) occurred between 3 and 30 days after the procedure.

### Clinical predictors of ischemic stroke

Results of univariate analysis for all potential clinical predictors are shown **in Supplemental Table 2.** Multivariable analysis identified several significant clinical predictors of ischemic stroke. Female sex (OR, 1.20), age > 85 years (OR, 1.25), weight < 60 kg (OR, 1.26), symptomatic status (OR, 1.85), previous stroke or transient ischemic attack (TIA) (OR, 3.19), more than one episode of acute heart failure (AHF) (OR, 1.20), severe mobility reduction (OR, 1.30), diabetes (OR, 1.17) and reduced creatinine clearance of < 60 mL/min (OR, 1.24) were associated with an increased risk of stroke **(Figure 2)**. **Supplemental Figure S1** summarizes the crude incidence of post-TAVI stroke according to the presence or absence of identified predictors.

**Figure 2.**
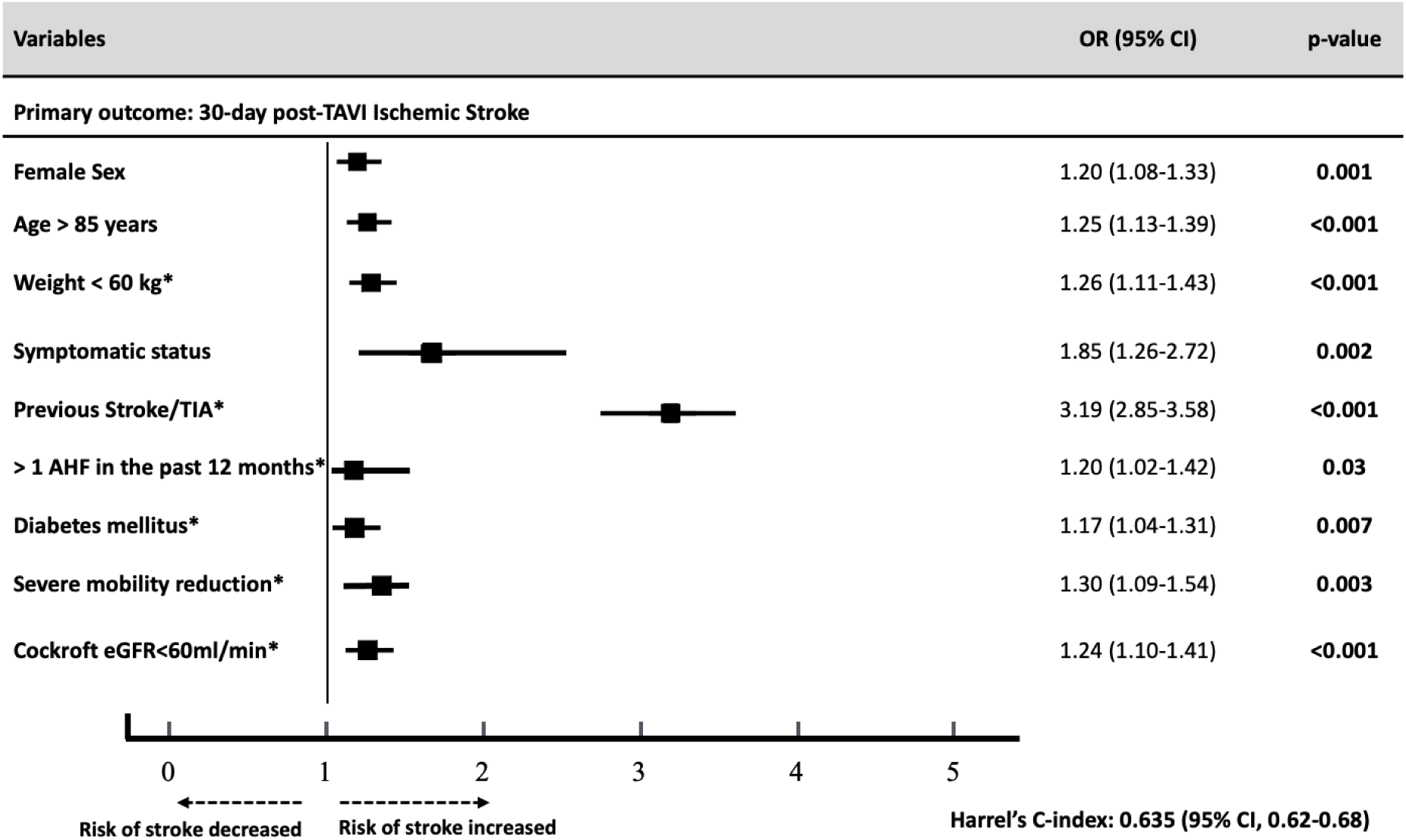
Multivariable Analysis of Factors Associated with Ischemic Stroke within 30 Days after Transcatheter Aortic Valve Implantation (TAVI) with femoral access. AHF: Acute Heart Failure; TIA: Transient Ischemic Attack; CKD: Chronic Kidney Disease. * Imputed variables.

### Risk-score construction

Points were assigned to variables to create a point-score model (range, 0-10, step size of 1) which we call the STRoke After Tavi (STRAT) Score **(Figure 3).** Higher risk scores were at greater risk for 30-day ischemic stroke; the odds ratio for stroke per one increment increase in the score was 1.38 (95% CI, 1.37-1.39, p ≤0.001) **(Figure 4).**

**Figure 3.**
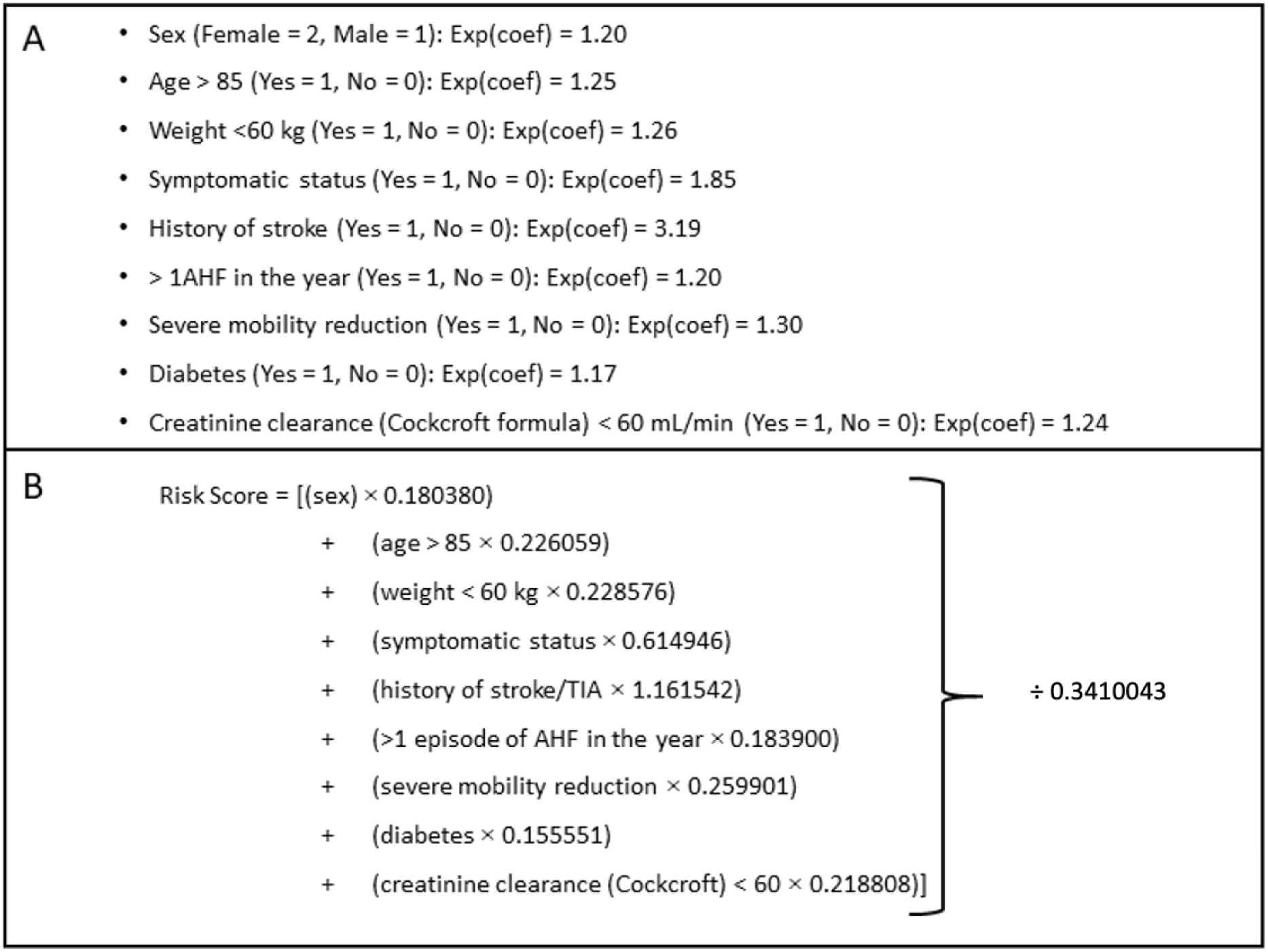
**A. Variables Included in the Risk Score and Their Modalities** **B. Risk Score Formula** The risk score was calculated using the following formula based on the β coefficients from the multivariable analysis. Each variable’s contribution to the risk score is determined by multiplying the presence (2), (1) or absence (0) of the characteristic by its corresponding β coefficient from the multivariable analysis. The final score was divided by a factor of 0,3410043 (sum of the beta coefficients/10) to make the score more easily interpretable, ranging between 0 and 10.

**Figure 4.**
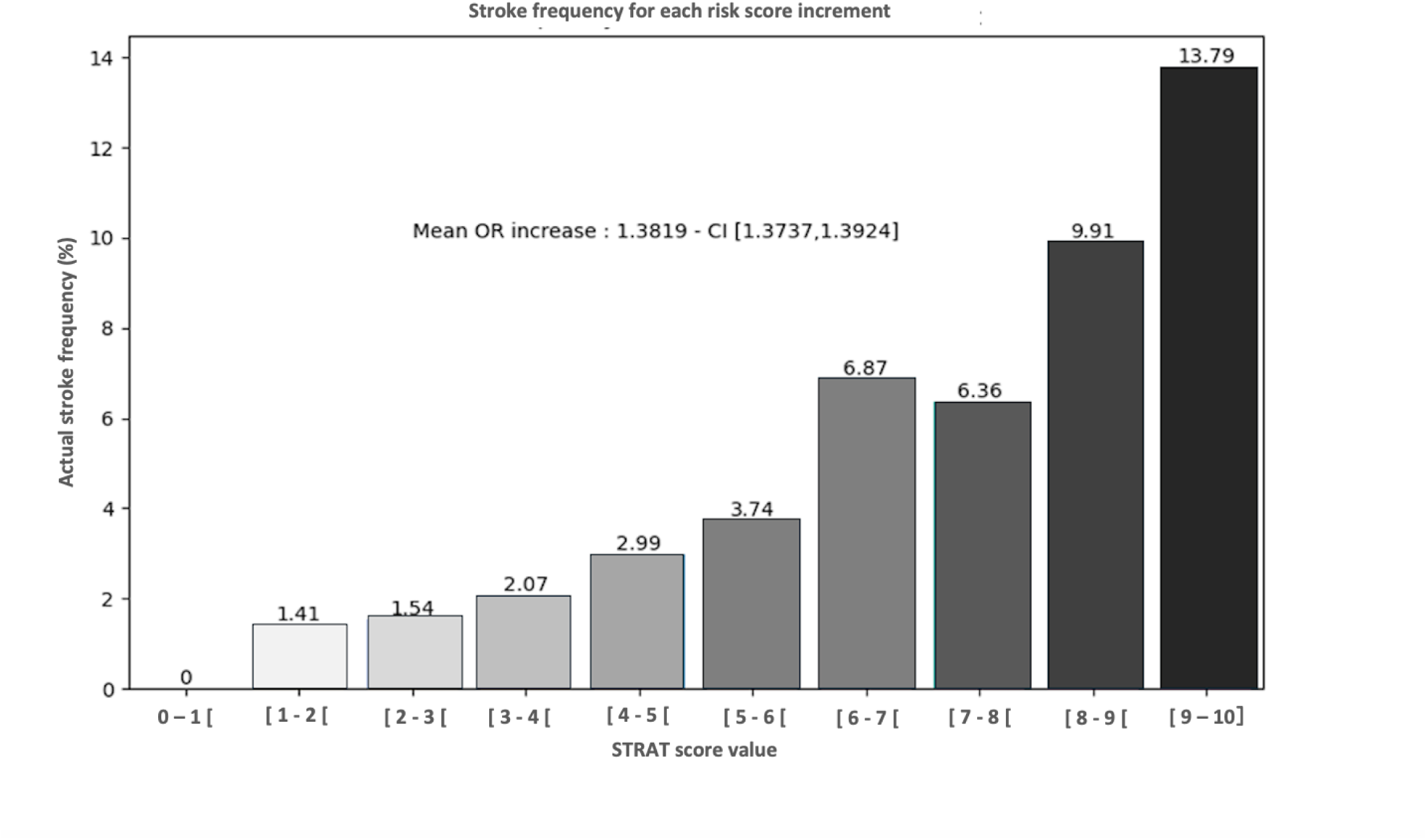
Observed rates of 30-day ischemic stroke (IS) according to the STRoke After TAVI (STRAT) score increments.

### Risk-score performance and internal validation

The Brier score of the STRAT Score was 0.178 (95% CI: 0.177-0.179) and the C-statistic was 0.63. In the visual plot comparing predicted and observed stroke rates **(Figure 5)**, the predicted rates closely matched the observed rates across all risk levels. Calibration-in-the-large, which assesses the overall agreement between predicted and observed probabilities, showed an intercept value of 0.014 (95% CI: 0.006-0.023; best value = 0) suggesting good overall calibration. The calibration slope, which quantifies the spread of estimated risks relative to observed outcomes, was found to be 0.78 (95% CI, 0.66-0.90; best value = 1), suggesting a slight tendency to underestimate the risk among the lowest and overestimate the risk among the highest risk patients (**Supplemental Figure S2)**.

**Figure 5.**
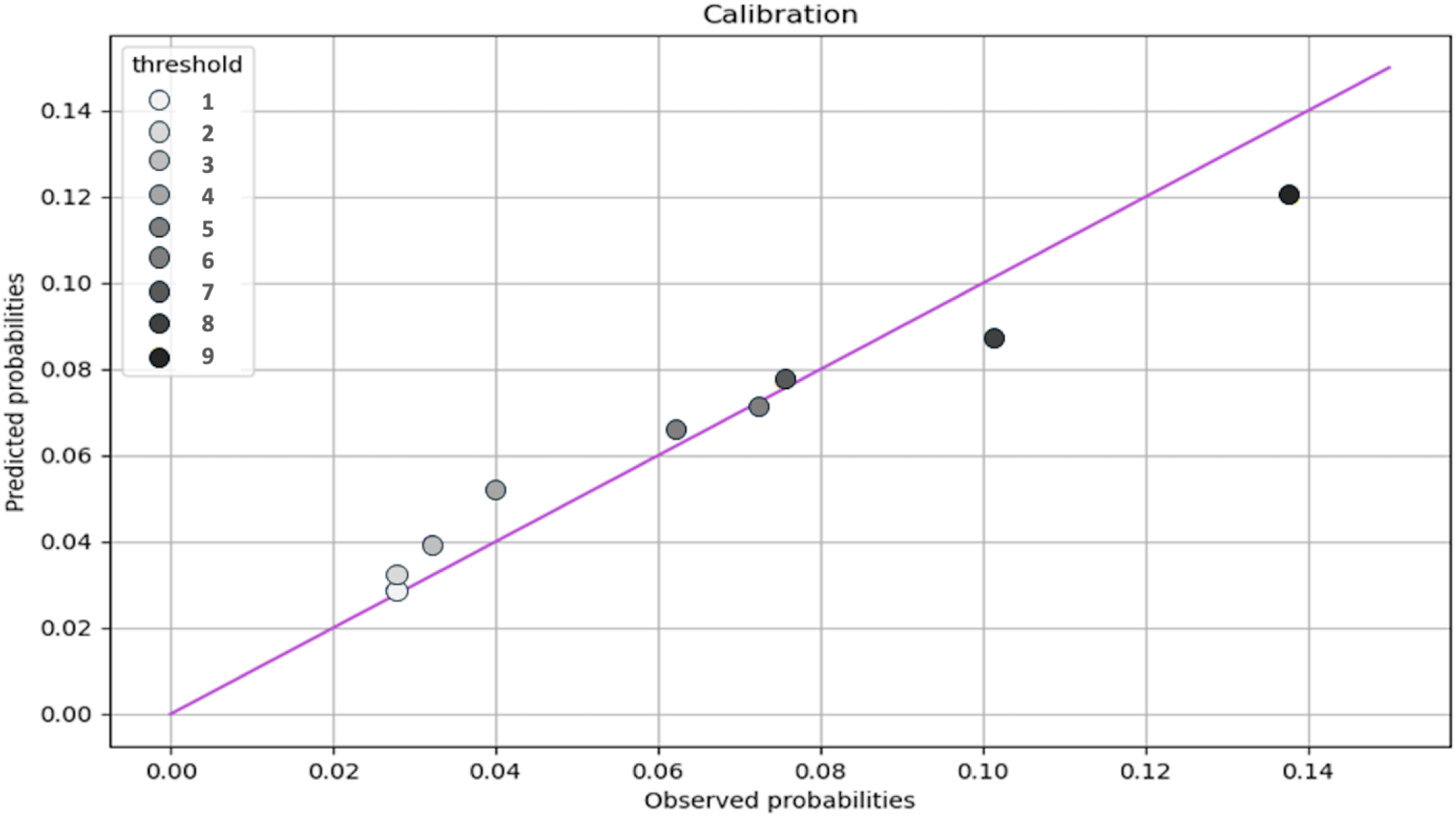
Calibration Curve for Predicted vs Observed Probabilities. The calibration plot compares the predicted probabilities of 30-day ischemic stroke (IS) from the model to the observed probabilities at every 1 score interval (with scores ranging from 0 to 10). Models are well calibrated if the predicted risks (circles) are close to the reference line.

The internal validity of the model was assessed by cross-validation. The internally validated Brier score was 0.1211, the C-statistic was 0.63 and calibration-in-the-large intercept 0.012.

### Risk-score predicted risk classification

Patients were classified into three risk categories for ischemic stroke, based on the total point scores and according to the appropriate calibration. The largest proportion of patients were classified as low risk (90.19%, 95% CI, 89.96%-90.44%), followed by intermediate risk (7.98%, 95% CI, 7.23%-8.73%) and high risk (1.83%, 95% CI, 1.05%-2.61%). The ischemic stroke frequency increased from 2.25% in the low-risk group (score <6) to 6.51% in the intermediate-risk group (6 ≤ score <8) and 10.10% in the high-risk group (score ≥8). The odds ratio for ischemic stroke in the intermediate-risk group (6 ≤ score <8) and high-risk group compared to the low-risk group was 3.05 (95% CI, 2.69-3.47, p <0.001) and 4.85 (95% CI, 3.93-5.98, p <0.001), respectively. The odds ratio for stroke between the high-risk and intermediate-risk groups was 1.59 (95% CI, 1.32-1.96) **(Figure 6)**. The cumulative incidence of ischemic strokes for the three groups, using Kalbfleisch and Prentice cumulative incidence curves (KPC), is presented in **Supplemental Figure S3.**

**Figure 6.**
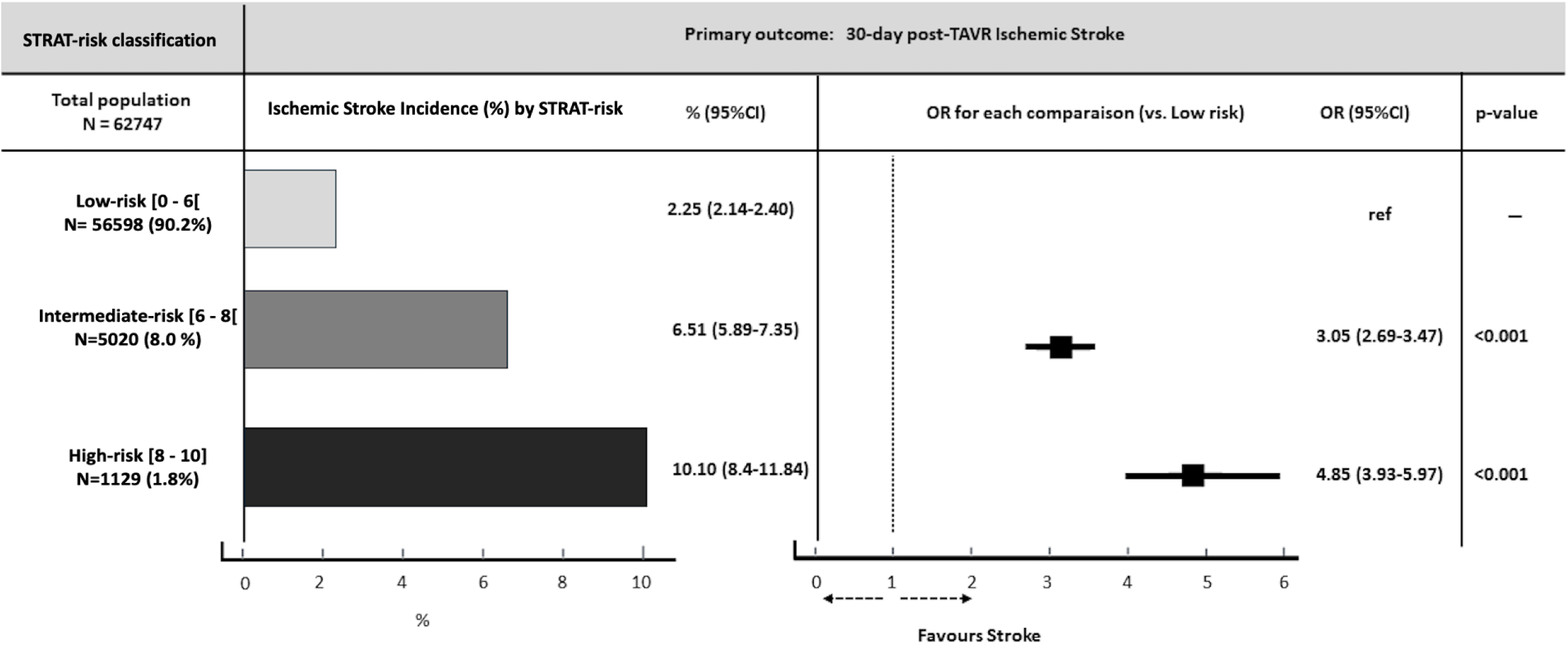
Performance of the STRAT score-defined risk categories for the prediction of 30-day stroke after TAVI. STRoke After Transcatheter Aortic Valve Implantation (STRAT) score risk staging increased with point totals: Low risk ([0, 6[), intermediate risk ([6, 8[), and high risk ([8, 10]). The odds ratios (OR) with their 95% confidence intervals (CI) are provided for comparison with the low-risk group.

### Sensitivity analysis

The STRAT score consistently identified higher odds ratios (ORs) for ischemic stroke across all predefined clinical subcategories, including complete cases, intervention periods (2013-2015, 2016-2018, 2019-2021), center intervention volumes (high vs. low), peri-procedural ischemic strokes (0-3 days), and ICD-10 code I63 (cerebral infarction) **(supplemental Figure S4).** We observed similar results in the internal-external validation performed by dividing the cohort into five major geographic regions of France **(supplemental Figure S5).** The Kaplan– Meier curves illustrate the 30-day mortality risk within each point score level of the STRAT score. Mortality rates ranged between 1.7 % and 14 %. There was no direct correspondence between each individual score level and increasing increments of mortality (Competing risk analysis) (**Supplemental Figure S6**).

## DISCUSSION

We derived a risk score fully dedicated to stroke prediction in patients undergoing TAVI, namely the STRAT score, based on easily accessible clinical parameters from a nationwide population of more than 60,000 participants. When collapsed into a 3-category risk score, this scoring system was able to identify a sizeable proportion of patients across clinically meaningful categories, namely low and intermediate/high risk of stroke (90.0% and 10.0% of the development population, respectively). The calculated discrimination capacity was moderate, and the associated calibration capacity was excellent, by categorizing patients according to relevant risk thresholds. These results suggest that the STRAT score identifies TAVI patients with a higher stroke risk, which could assist providers in potentially adjusting management.

The STRAT score includes several patient-related stroke risk factors that were previously found to be associated with an increased risk of cerebrovascular event after TAVI, namely higher age, previous history of stroke, female sex, diabetes, chronic kidney disease, and low body weight [24, 25]. Multiple other predictors of stroke have been described, including clinical (e.g. atrial fibrillation[25]), pre-procedural (e.g. bicuspid valve [26], aortic wall thrombus [27]) or procedural features (e.g. procedure time [28]). However, when considered independently, these different predictors cannot achieve a predicted probability of stroke beyond 3.0% each. Furthermore, the post-TAVI stroke rate has remained stable over time in real-life registries, at approximately 2.5%, despite technological improvements and increased operator experience. This suggests that there may be an incompressible baseline risk in the overall population undergoing TAVI [8]. This baseline stroke rate was reflected in the lowest risk category of our risk score, which comprised 90% of the development cohort.

The risk stratification achieved by the STRAT score indicated that the remaining 10% of our population faced a higher stroke risk, with a three-fold increase in risk in the intermediate category, and an almost five-fold increase in risk in those classed as high risk. No such findings have ever been documented with contemporary TAVI techniques [8]. Identifying TAVI patients who have an intermediate/ high risk of stroke may prompt use of risk-guided preventive strategies [29]. In this context, cerebral embolic protection (CEP) devices have shown significant safety and efficacy in capturing stroke-causing debris during TAVI, with high procedural success rates and minimal adverse events [30,31]. Using the STRAT score could help to identify patients in whom CEP might be warranted.

A certain proportion of patients either die or experience no improvement in quality-of-life shortly after TAVI, despite randomized controlled trials showing that TAVI improves survival and symptoms compared to control strategies [6, 32]. This underscores the need to identify candidates for whom a TAVI procedure may be futile. A high-risk classification by our scoring system (corresponding to 1.8% of patients with a stroke risk >10%) could prompt reconsideration of TAVI. Indeed, in cases where the expected benefits are already uncertain, especially if a cerebral protection device is not feasible due to anatomical constraints, the risk assessment could provide an additional argument to inform decision-making. This approach would help refine the risk-benefit ratio of TAVI, potentially avoiding futile interventions [33].

This study has some limitations. Firstly, we had a limited panel of variables determined by the case report form from FRANCE TAVI. Specifically, we did not have certain computed tomography imaging parameters known to be potential predictors of stroke in the TAVI population (e.g. hypo attenuated leaflet thickening, atherosclerotic burden in the proximal aorta) [8]. Additionally, data concerning antithrombotic treatment during and immediately after the procedure were lacking. Finally, we dichotomized variables to facilitate creation of a risk score, but this may provide less granular information than when continuous variables are used.

### Conclusion

The STRAT score is the first clinical predictive model to identify patients at increased risk of ischemic stroke post-TAVI, using the extensive nationwide FRANCE TAVI registry. Further studies are warranted to externally validate the score and determine its clinical usefulness in the setting of stroke prevention.

## Data Availability

The data supporting the findings of this study are available within the article and its supplementary materials. Additional data may be available from the corresponding author upon reasonable request, in accordance with national data protection regulations and FRANCE-TAVI registry policies.

## Disclosures of interest

N Meneveau declares consultancy fees from Edwards, speaker fees from Medtronic, and grants from Medtronic.

G. Cayla has received research grants/consultant fees/lectures fees from Edwards and Medtronic.

No other author has anything to declare.

## Ethical approval

All patients provided written informed consent for anonymous processing of their data, and the Institutional Review Board of the French Ministry of Health approved the registry.

## Pre-registered clinical trial number

Not applicable

---

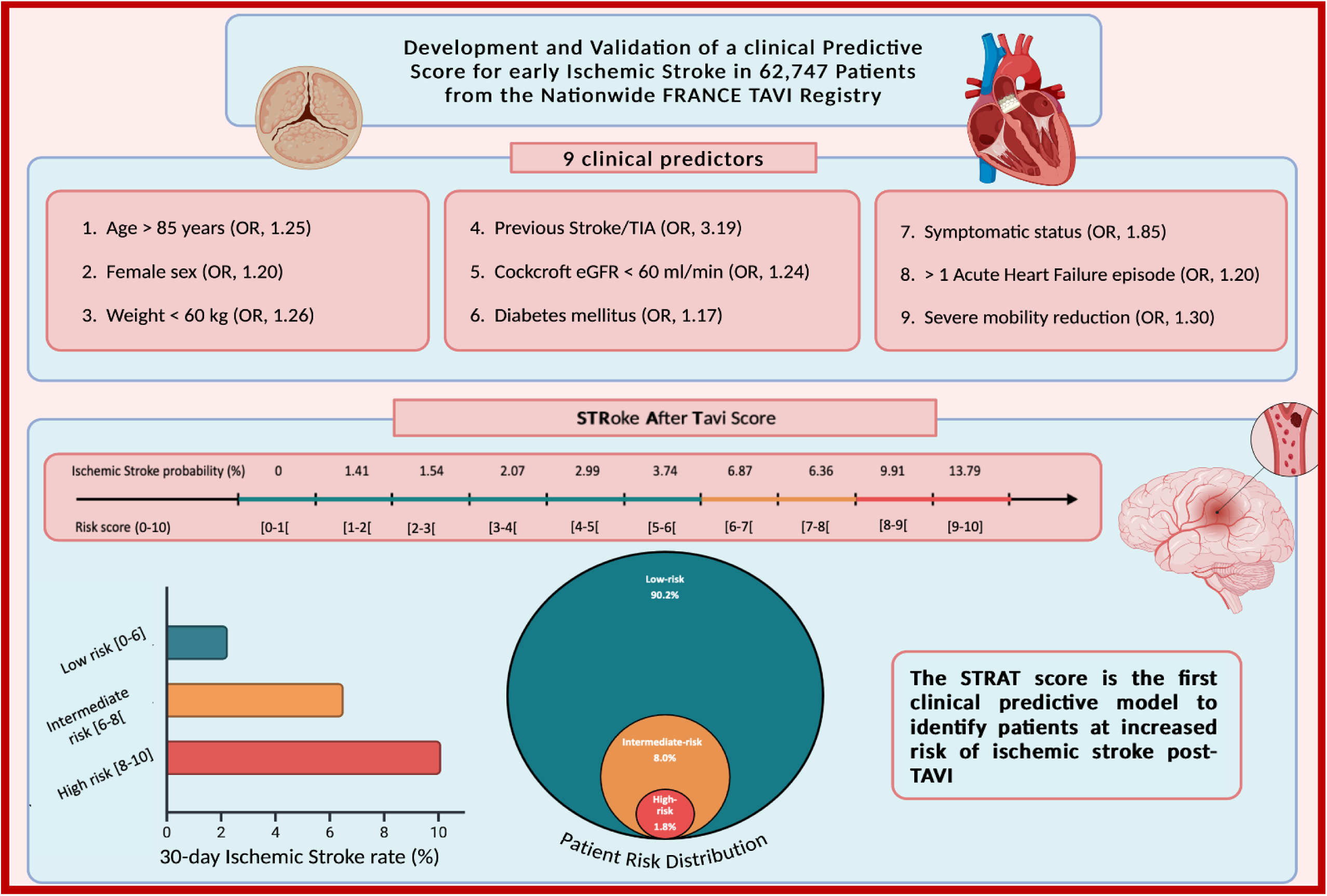

## References

1. Leon, M.B., et al., Transcatheter Aortic-Valve Implantation for Aortic Stenosis in Patients Who Cannot Undergo Surgery. New England Journal of Medicine, 2010. 363(17): p. 1597–1607.

2. Leon, M.B., et al., Transcatheter or Surgical Aortic-Valve Replacement in Intermediate-Risk Patients. New England Journal of Medicine, 2016. 374(17): p. 1609–1620.

3. Smith, C.R., et al., Transcatheter versus surgical aortic-valve replacement in high-risk patients. N Engl J Med, 2011. 364(23): p. 2187–98.

4. Adams, D.H., et al., Transcatheter Aortic-Valve Replacement with a Self-Expanding Prosthesis. New England Journal of Medicine, 2014. 370(19): p. 1790–1798.

5. Vahanian, A., et al., 2021 ESC/EACTS Guidelines for the management of valvular heart disease. Eur Heart J, 2022. 43(7): p. 561–632.

6. Otto, C.M., et al., 2020 ACC/AHA Guideline for the Management of Patients With Valvular Heart Disease: A Report of the American College of Cardiology/American Heart Association Joint Committee on Clinical Practice Guidelines. Circulation, 2021. 143(5): p. e72–e227.

7. Carroll, J.D., et al., STS-ACC TVT Registry of Transcatheter Aortic Valve Replacement. J Am Coll Cardiol, 2020. 76(21): p. 2492–2516.

8. Reddy, P., et al., Cerebrovascular events after transcatheter aortic valve implantation. EuroIntervention, 2024. 20(13): p. e793–e805.

9. Tchetche, D., et al., Cerebrovascular Events Post-Transcatheter Aortic Valve Replacement in a Large Cohort of Patients. JACC: Cardiovascular Interventions, 2014. 7(10): p. 1138–1145.

10. Daniel, K., et al., What are the social consequences of stroke for working-aged adults? A systematic review. Stroke, 2009. 40(6): p. e431–40.

11. Lai, S.M., et al., Persisting consequences of stroke measured by the Stroke Impact Scale. Stroke, 2002. 33(7): p. 1840–4.

12. Miller, D.C., et al., Transcatheter (TAVR) versus surgical (AVR) aortic valve replacement: occurrence, hazard, risk factors, and consequences of neurologic events in the PARTNER trial. J Thorac Cardiovasc Surg, 2012. 143(4): p. 832–843 e13.

13. Auffret, V., et al., Temporal Trends in Transcatheter Aortic Valve Replacement in France: FRANCE 2 to FRANCE TAVI. J Am Coll Cardiol, 2017. 70(1): p. 42–55.

14. Didier, R., et al., Successful linkage of French large-scale national registry populations to national reimbursement data: Improved data completeness and minimized loss to follow-up. Arch Cardiovasc Dis, 2020. 113(8-9): p. 534–541.

15. Collins, G.S., et al., Evaluation of clinical prediction models (part 1): from development to external validation. BMJ, 2024. 384: p. e074819.

16. Barnard, J. and X.L. Meng, Applications of multiple imputation in medical studies: from AIDS to NHANES. Stat Methods Med Res, 1999. 8(1): p. 17–36.

17. Woldendorp, K., et al., Silent brain infarcts and early cognitive outcomes after transcatheter aortic valve implantation: a systematic review and meta-analysis. Eur Heart J, 2021. 42(10): p. 1004–1015.

18. Perkins, N.J., et al., Principled Approaches to Missing Data in Epidemiologic Studies. Am J Epidemiol, 2018. 187(3): p. 568–575.

19. Johnston, R., K. Jones, and D. Manley, Confounding and collinearity in regression analysis: a cautionary tale and an alternative procedure, illustrated by studies of British voting behaviour. Qual Quant, 2018. 52(4): p. 1957–1976.

20. Mehta, H.B., et al., Regression coefficient-based scoring system should be used to assign weights to the risk index. J Clin Epidemiol, 2016. 79: p. 22–28.

21. Austin, P.C., D.S. Lee, and J.P. Fine, Introduction to the Analysis of Survival Data in the Presence of Competing Risks. Circulation, 2016. 133(6): p. 601–9.

22. Collins, G.S., et al., Transparent Reporting of a Multivariable Prediction Model for Individual Prognosis or Diagnosis (TRIPOD): The TRIPOD Statement. Eur Urol, 2015. 67(6): p. 1142–1151.

23. Cuschieri, S., The STROBE guidelines. Saudi J Anaesth, 2019. 13(Suppl 1): p. S31–S34.

24. Vlastra, W., et al., Predictors, Incidence, and Outcomes of Patients Undergoing Transfemoral Transcatheter Aortic Valve Implantation Complicated by Stroke. Circ Cardiovasc Interv, 2019. 12(3): p. e007546.

25. Auffret, V., et al., Predictors of Early Cerebrovascular Events in Patients With Aortic Stenosis Undergoing Transcatheter Aortic Valve Replacement. Journal of the American College of Cardiology, 2016. 68(7): p. 673–684.

26. Makkar, R.R., et al., Association Between Transcatheter Aortic Valve Replacement for Bicuspid vs Tricuspid Aortic Stenosis and Mortality or Stroke Among Patients at Low Surgical Risk. JAMA, 2021. 326(11): p. 1034–1044.

27. Bonnet, M., et al., Association Between Aortic Wall Thrombus and Thromboembolic Events After Transfemoral Transcatheter Aortic Valve Replacement. JACC Cardiovasc Interv, 2024. 17(14): p. 1680–1690.

28. De Backer, O., et al., Early and late risk of ischemic stroke after TAVR as compared to a nationwide background population. Clin Res Cardiol, 2020. 109(7): p. 791–801.

29. Khera, S., et al., Prevention and Management of Stroke After Transcatheter Aortic Valve Replacement: The Mount Sinai Stroke Initiative. J Am Heart Assoc, 2023. 12(4): p. e028182.

30. Wolfrum, M., et al., Cerebral embolic protection during transcatheter aortic valve replacement: a systematic review and meta-analysis of propensity score matched and randomized controlled trials using the Sentinel cerebral embolic protection device. BMC Cardiovasc Disord, 2023. 23(1): p. 306.

31. Kapadia, S.R., et al., Cerebral Embolic Protection during Transcatheter Aortic-Valve Replacement. New England Journal of Medicine, 2022. 387(14): p. 1253–1263.

32. Hindricks, G., et al., 2020 ESC Guidelines for the diagnosis and management of atrial fibrillation developed in collaboration with the European Association of Cardio-Thoracic Surgery (EACTS). Eur Heart J, 2020.

33. Lindman, B.R., et al., Futility, benefit, and transcatheter aortic valve replacement. JACC Cardiovasc Interv, 2014. 7(7): p. 707–16.

